# Machine learning and data-driven models for predicting post-stroke dysphagia: a systematic review and meta-analysis

**DOI:** 10.64898/2026.07.15.26358113

**Authors:** Soheil Mohammadi Yazdi, Mohsen Motevaselian, Seyed Sepehr Khatami, Nazli Radfar, Zahra Jourahmad, Hugo Andres Perez

**Affiliations:** Department of Neurology, Saint Louis University School of Medicine, St. Louis, Missouri, USA; Department of Radiology and Imaging Sciences, Emory University School of Medicine, Atlanta, Georgia, USA; Department of Neurology, Loma Linda University Health, Loma Linda, California, USA; Department of Neurology, Penn State College of Medicine, Hershey, Pennsylvania, USA; Department of Neurosurgery, Baylor College of Medicine, Houston, Texas, USA; Sanavate Hospital, Houston, Texas, USA

**Keywords:** Post-stroke dysphagia, Stroke, Deglutition disorders, Machine learning, Clinical prediction model, Area under the curve, Meta-analysis

## Abstract

**Background:** Post-stroke dysphagia (PSD) contributes to aspiration, pneumonia, malnutrition, prolonged hospitalization and mortality. We evaluated the discrimination, validity and readiness of machine learning and data-driven prediction models for PSD-related outcomes.

**Methods:** Following a prospectively registered protocol (PROSPERO CRD420261419259), we searched PubMed/MEDLINE, Embase, Web of Science Core Collection, CINAHL and CENTRAL from inception through June 7, 2026. Eligible studies developed or validated multivariable prediction models for PSD-related outcomes in adults with stroke. We used PROBAST and PROBAST+AI to assess risk of bias and applicability and TRIPOD+AI to evaluate reporting. Area under the curve (AUC) estimates were pooled on the logit scale with random-effects models.

**Results:** Twenty-four studies were included and ten contributed to meta-analysis. Four studies predicting early or incident PSD yielded a pooled AUC of 0.94 (95% CI 0.60–0.99; I² = 95.6%). Pooled AUCs were 0.84 (95% CI 0.71–0.92) for aspiration or penetration-aspiration and 0.89 (95% CI 0.24–1.00) for severe dysphagia. The exploratory analysis of all ten risk-prediction models produced an AUC of 0.90 (95% CI 0.80–0.95), but heterogeneity was substantial (I² = 90.3%) and the prediction interval was 0.51–0.99. Every study had high risk of bias because of analysis-domain concerns; calibration and external validation were uncommon.

**Conclusions:** Reported discrimination was often high, but the evidence does not establish reliable performance in care. Independent validation, calibration, complete model reporting and clinical-impact studies are needed before these models guide post-stroke swallowing care.

## 1. Introduction

Post-stroke dysphagia (PSD) is an impairment of swallowing following stroke, most commonly affecting the oral and pharyngeal phases and potentially leading to penetration or aspiration.[1] Its reported frequency varies according to the method of assessment. PSD affects approximately 40–47% of patients with acute stroke, and a recent meta-analysis of 34 cohorts reported an overall prevalence of 46.6%. Reported prevalence is generally lower with bedside screening, at approximately 37–45%, and higher with instrumental assessment, at approximately 64–78%, partly because instrumental testing can detect clinically occult swallowing impairment.[1–3] PSD is associated with aspiration, stroke-associated pneumonia, malnutrition, dehydration, prolonged hospitalization, poorer functional recovery and increased mortality.[2,4–5]

Early recognition allows timely implementation of measures aimed at reducing aspiration and nutritional complications. Current guidelines therefore recommend swallowing screening for all patients with acute stroke before oral intake, including oral medication.[6] Screening may be performed by speech-language pathologists or other appropriately trained healthcare professionals. Instrumental assessment with fiber-optic endoscopic evaluation of swallowing (FEES) or videofluoroscopic swallowing study (VFSS) may be used when aspiration is suspected, bedside findings are inconclusive, or a more detailed assessment of swallowing physiology and severity is required.[6–7] The diagnostic accuracy of bedside screening tools varies across instruments and reference standards, while universal instrumental assessment is limited by cost, availability and the need for trained personnel. These limitations have created interest in point-of-care risk-stratification tools that could complement, rather than replace, routine swallowing screening.[7–8] Differences in diagnostic criteria and reference standards across studies further complicate comparison of reported estimates.[2]

Several distinctions are important when evaluating prediction models in this field. Penetration and aspiration refer to entry of material into the laryngeal airway and are commonly graded using the Penetration-Aspiration Scale; they represent specific manifestations of swallowing impairment rather than dysphagia as a whole.[9] Swallowing function improves within the first weeks in many patients, while clinically important dysphagia persists in a substantial minority. Accordingly, models predicting the occurrence of PSD address a different clinical question from prognostic models predicting recovery, persistence or feeding-tube dependence.[10] Frequently reported predictors across these tasks include older age, greater stroke severity, previous stroke, and selected stroke subtypes or lesion locations.[3,10]

Machine-learning methods can model nonlinear associations and interactions among multiple predictors and have increasingly been applied to stroke outcomes, including dysphagia and aspiration prediction.[11–13] However, greater methodological flexibility does not necessarily translate into superior clinical performance compared with well-developed regression models. Previous reviews have described this literature without quantitatively pooling estimates of model discrimination. A 2026 scoping review identified 38 studies of intelligent technologies for dysphagia prediction across several high-risk populations, including stroke, but did not perform a meta-analysis.[12] A systematic review of aspiration-risk prediction models in stroke summarized 18 studies without pooling discrimination estimates.[13] A separate review synthesized clinical predictors of dysphagia recovery but did not meta-analyze the discrimination of multivariable prediction models.[10] To our knowledge, no previous meta-analysis has pooled the discrimination of machine-learning and other data-driven prediction models for PSD-related outcomes.

We therefore aimed to identify and critically appraise machine-learning and other data-driven multivariable prediction models for PSD-related outcomes in adults with stroke. When studies were sufficiently comparable, we pooled area under the receiver operating characteristic curve and C-statistic estimates, specifying early or incident PSD as the primary outcome. Finally, we assessed risk of bias and applicability using PROBAST and PROBAST+AI, evaluated reporting against TRIPOD+AI, and determined the extent to which the available evidence supports clinical implementation.[14–16]

## 2. Methods

### 2.1 Protocol and registration

This systematic review was reported in accordance with the PRISMA 2020 statement and was prospectively registered in PROSPERO (CRD420261419259).[17] The registered protocol is available through PROSPERO under that identifier. The eligibility criteria, search strategy, data items, risk-of-bias assessment and synthesis plan were specified before study selection began. Deviations from the registered protocol are described in Section 2.7.

### 2.2 Eligibility criteria

Eligible studies developed, validated, updated or compared machine-learning or other explicitly data-driven multivariable prediction models for PSD-related outcomes in adults aged 18 years or older with ischemic stroke, intracerebral hemorrhage, subarachnoid hemorrhage, cerebral venous thrombosis associated with stroke, or mixed adult stroke cohorts. Mixed cohorts were eligible when stroke-specific results were extractable or patients with stroke constituted the clear majority of the study population.

Eligible models included machine-learning and deep-learning algorithms, neural networks, tree-based ensemble methods and support vector machines, as well as data-driven multivariable models derived using methods such as least absolute shrinkage and selection operator (LASSO), elastic net and nomogram-based modeling. Conventional regression models were eligible when they were developed or validated for individualized outcome prediction and reported extractable measures of model performance.

The primary outcome was early or incident PSD. Secondary outcomes included any PSD, aspiration or penetration-aspiration, severe dysphagia, persistent or prolonged dysphagia, swallowing recovery, feeding-tube dependence and automated interpretation of instrumental or physiological assessments, including VFSS, FEES, acoustic assessment and ultrasound.

We excluded studies involving pediatric, animal or non-stroke populations; univariable analyses or risk-factor studies that did not develop or validate a multivariable prediction model; treatment-effect studies; reviews, protocols and editorials; and studies without extractable model-performance data.

### 2.3 Information sources and search strategy

We searched five databases from inception through June 7, 2026, without language restrictions: PubMed, Embase, Web of Science Core Collection, CINAHL via EBSCO and the Cochrane Central Register of Controlled Trials (CENTRAL). The reference lists of relevant articles were also screened to identify additional eligible studies. Complete database-specific search strategies are provided in Supplementary Methods S1.

### 2.4 Study selection and data extraction

Records were deduplicated sequentially using normalized digital object identifiers, PubMed identifiers and exact normalized titles. Screening and data extraction were conducted using a predefined codebook. Uncertain records, all included studies and quantitative data used in the analyses were checked by a second reviewer, and disagreements were resolved by consensus.

Data extraction was guided by the CHARMS framework and included study design, population characteristics, stroke type and phase, sample size and number of outcome events, outcome definition and reference standard, model class and algorithm, predictor modality and timing, validation design, discrimination, calibration, and availability of code and data.[18] Unreported or unclear items were recorded as such; no study characteristics were imputed.

When a study reported multiple models, one performance estimate was selected per cohort. Preference was given to the authors’ final or recommended model or, when necessary, to the model with the strongest validation design and clinically feasible predictors. Performance from external, temporal or geographic validation was prioritized over performance obtained through resampling, split-sample validation or evaluation in the development dataset. Overlapping cohorts were not counted more than once.

### 2.5 Risk of bias, applicability and reporting completeness

Risk of bias and applicability were assessed using PROBAST. Machine learning models were additionally appraised using PROBAST+AI across the participant, predictor, outcome and analysis domains.[14–15] Reporting completeness was evaluated against the TRIPOD+AI statement.[16] Prediction-model-specific frameworks were used instead of generic quality-assessment tools, in accordance with recommendations for systematic reviews of prognostic and diagnostic prediction models.[19] We did not apply GRADE or another formal certainty framework because the review synthesized prediction-model performance rather than intervention effects. Confidence in the evidence was considered through risk of bias, applicability, validation design, calibration, heterogeneity and reporting completeness.

### 2.6 Statistical analysis

The area under the receiver operating characteristic curve, reported as the AUC or C-statistic, was the primary measure of model discrimination.[20] AUC estimates were transformed to the logit scale before pooling and were subsequently back-transformed for presentation.

When a study reported a 95% confidence interval for the AUC, the standard error of the logit-transformed AUC was derived from the transformed confidence limits. When a confidence interval was unavailable, the variance was approximated using the Hanley–McNeil method based on the numbers of participants with and without the outcome, and these estimates were flagged accordingly.[20] Confidence limits reaching 0 or 1 were identified as boundary values because they could not be directly transformed on the logit scale. Study-level inputs and variance sources are provided in Supplementary Table S3.

The prespecified primary analysis pooled AUC estimates from models predicting early or incident PSD. Separate secondary analyses pooled models predicting aspiration or penetration-aspiration and severe dysphagia. As a secondary exploratory analysis, all eligible clinical risk-prediction models were also pooled regardless of the specific PSD-related outcome. This overall analysis was intended to describe reported discrimination across data-driven models for post-stroke swallowing complications and was not interpreted as evidence that the different clinical outcomes were interchangeable.

Random-effects meta-analysis was performed when at least three studies were considered sufficiently comparable. Between-study variance was estimated using restricted maximum likelihood, and confidence intervals were calculated using the Hartung–Knapp adjustment.[21–22] Statistical heterogeneity was described using τ², I² and Cochran’s Q statistic. A 95% prediction interval was also reported when feasible.[23–24] Forest plots displayed study-specific and pooled AUCs with 95% confidence intervals, grouped by outcome stratum. Sensitivity analyses excluded boundary-level estimates and estimates whose variance required approximation; leave-one-out analyses were performed for the exploratory overall synthesis.

Subgroup analyses were interpreted descriptively because of the small number of studies within each category. Small-study effects were not formally evaluated because only ten clinically heterogeneous studies were available; funnel plots and formal tests are unreliable in this setting.[25] Decision-curve analysis and net benefit were not pooled because they were infrequently reported and depend on specific risk thresholds, clinical decisions and the relative consequences of false-positive and false-negative classifications.[26] Statistical analyses were performed in R using the metafor package.[27]

### 2.7 Deviations from the registered protocol

Three deviations from the registered protocol are reported for transparency. First, in addition to the prespecified primary analysis of early or incident PSD, we conducted a post hoc exploratory analysis pooling all eligible clinical risk-prediction models across outcome categories. Because this analysis combined different clinical targets, it was treated as a broad summary of reported discrimination and not as a clinically transportable estimate for any single PSD-related outcome.

Second, several prespecified subgroup and sensitivity analyses could not be performed as planned because of the limited number of eligible studies. In particular, the planned analysis excluding studies at high risk of bias in the analysis domain could not be conducted because every included study was rated as high risk in that domain. Analyses were therefore reported only when the available number of studies permitted meaningful evaluation.

Third, the author group was expanded from the four investigators listed in the PROSPERO registration to include two additional contributors who met the criteria for authorship. The eligibility criteria, search strategy, outcome definitions and risk-of-bias approach remained unchanged.

## 3. Results

### 3.1 Study selection

The search identified 1,973 records: 118 from PubMed, 164 from Embase, 111 from Web of Science, 1,548 from CINAHL via EBSCO, and 32 from Cochrane CENTRAL. After removal of 957 duplicates, 1,016 unique records underwent title and abstract screening. Seventy-eight reports, including 77 identified through database searching and one through citation searching, were sought for retrieval.

Three reports could not be retrieved, leaving 75 reports for full-text assessment. Fifty-one reports were excluded, most commonly because they did not include a multivariable prediction model (n = 13), involved a non-adult or non-stroke population (n = 10), did not report a swallowing-related outcome (n = 6), or evaluated an outcome unrelated to PSD (n = 6). Twenty-four studies met the inclusion criteria; ten contributed to the meta-analysis and 14 to the narrative or secondary synthesis (Figure 1; Supplementary Table S4).

**Figure 1.**
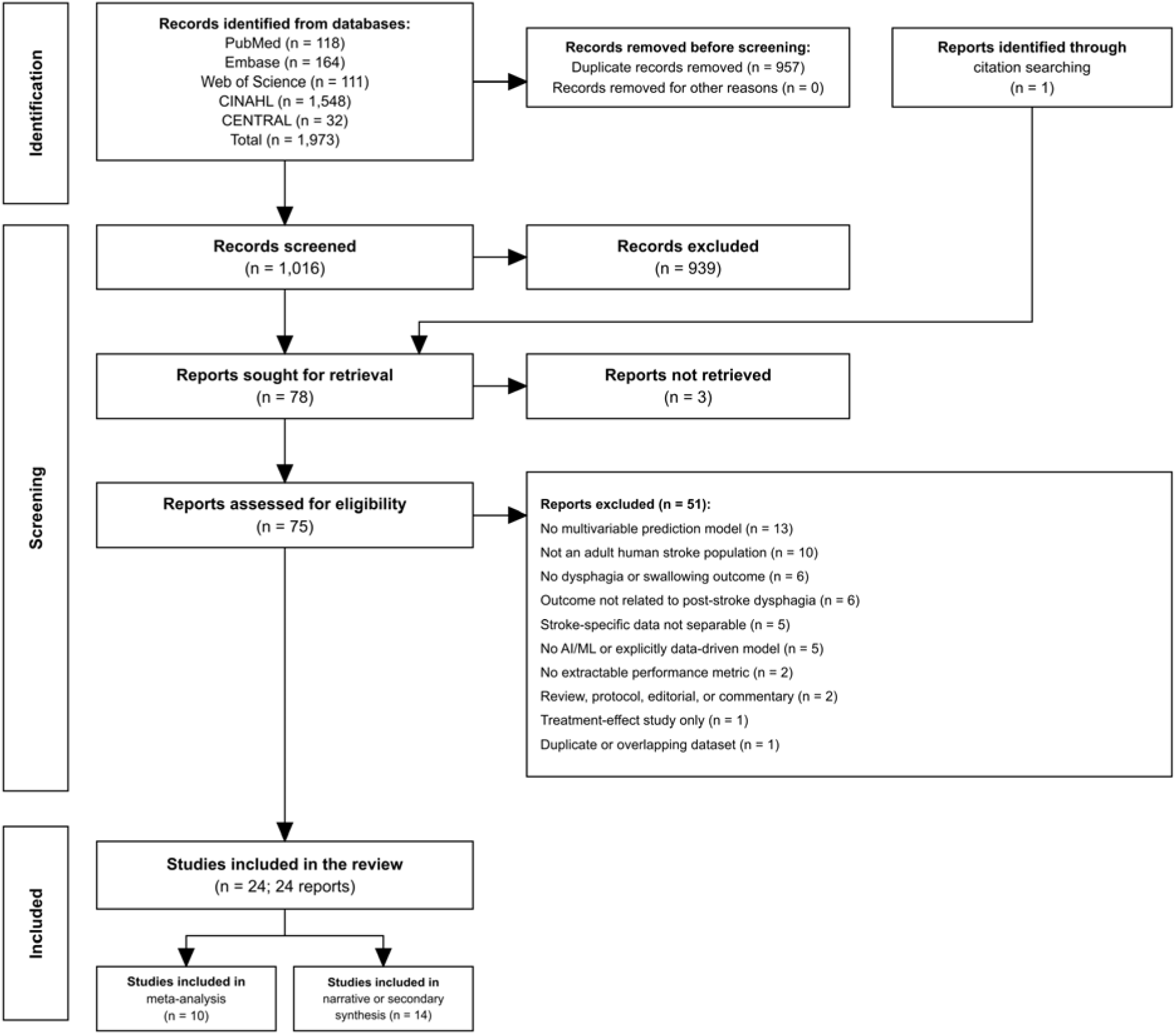
PRISMA 2020 flow diagram of study identification and selection. Of 1,973 database records and one report identified through citation searching, 78 reports were sought for retrieval, three were not retrieved, 75 were assessed for eligibility and 24 studies were included: ten in the meta-analysis and 14 in the narrative or secondary synthesis.

### 3.2 Study characteristics

The included studies were published between 2020 and 2026 and were conducted predominantly in China and the Republic of Korea, with additional studies from Japan, Canada and the United States. Sample sizes ranged from 68 to 3,408 participants.

Predictors were derived from clinical or electronic health record data, imaging, voice and acoustic signals, instrumental swallowing assessments, including VFSS and FEES, ultrasound, and multimodal combinations. The evaluated models included machine-learning algorithms, data-driven regression models or nomograms, and deep-learning approaches. Table 1 summarizes the ten studies included in the meta-analysis, and Table 2 summarizes the 14 studies included in the narrative or secondary synthesis.

**Table 1.** Characteristics of the ten meta-analyzed studies.

| Study | Country | Design; stroke type, phase | N (events) | Outcome stratum | Model — best (class) | Validation | AUC (95% CI) |
| --- | --- | --- | --- | --- | --- | --- | --- |
| Park 2023 [45] | Rep. Korea | Retrospective cohort; mixed, acute | 3,408 (448) | Aspiration/PA | Ridge regression (ML) | Internal split | 0.81 (0.76–0.86) |
| Ye 2024 [46] | China | Retrospective cohort; ischemic, acute | 724 (308) | Severe dysphagia | XGBoost (ML) | External cohort | 0.96 (0.93–0.99) |
| Wang 2024 [47] | China | Prospective cohort; ischemic, acute | 528 (89) | Aspiration/PA | LASSO nomogram (Reg) | Geographic external | 0.87 (0.78–0.96) |
| Chen 2024 [48] | China | Retrospective cohort; mixed, acute | 412 (184) | Aspiration/PA | LASSO nomogram (Reg) | Temporal | 0.88 (0.81–0.95) |
| Li 2025 [49] | China | Cross-sectional; ischemic, acute | 1,041 (305) | Early/incident PSD | Random forest (ML) | Internal split | 0.95 (0.93–0.98) |
| Lu 2025 [50] | China | Case-control; mixed, acute | 250 (100) | Early/incident PSD | Logistic nomogram (Reg) | Temporal† | 0.995 (0.98–1.00) |
| Sung 2026 [51] | Rep. Korea | Prospective cohort; ischemic, acute | 212 (66) | Severe dysphagia | Ultrasound-clinical score (Reg) | Internal split | 0.94 (0.87–1.00) |
| Zhang 2026 [52] | China | Retrospective cohort; ischemic, acute | 908 (464) | Early/incident PSD | Random forest (ML) | Internal split | 0.87 (0.83–0.91) |
| Weng 2026 [53] | China | Retrospective cohort; RSSI, acute | 394 (79) | Early/incident PSD | Logistic nomogram (Reg) | Bootstrap | 0.85 (0.80–0.90) |
| Lee 2026 [54] | Rep. Korea | Retrospective cohort; LMI, acute | 163 (44) | Severe dysphagia | Hier-ViT, imaging (DL) | Internal split | 0.69 (0.61–0.78) |

All ten studies were at high overall risk of bias (PROBAST/PROBAST+AI). †Confidence limit at a boundary ( $AUC \geq 0.99$ ); logit-AUC variance approximated by the Hanley–McNeil method. N (events) = total sample (outcome events). PA, penetration-aspiration; RSSI, recent small subcortical infarct; LMI, lateral medullary infarction; ML, machine learning; Reg, data-driven regression/nomogram; DL, deep learning; Hier-ViT, hierarchical vision transformer.

**Table 2.** Studies included in narrative or secondary synthesis (not meta-analyzed).

| Study | Country | Synthesis group | Outcome | Model — best (class) | Validation | AUC / metric |
| --- | --- | --- | --- | --- | --- | --- |
| Lee 2020 [55] | Rep. Korea | Prognostic / recovery | 6-month swallowing recovery | Bayesian network (ML) | Internal split | 0.80 |
| Tian 2026 [56] | China | Prognostic / recovery | Swallowing recovery | Gradient boosting (ML) | Temporal | 0.94 |
| Ito 2021 [57] | Japan | Prognostic / recovery | Prolonged dysphagia at discharge | Scoring system (Reg) | Internal split | 0.92 |
| Zhang 2022 [58] | China | Prognostic / recovery | Nasogastric-tube need at 1 week | Logistic nomogram (Reg) | Temporal | 0.79 |
| Seo 2025 [59] | Rep. Korea | Prognostic / recovery | Long-term PSD prognosis | Ensemble (ML) | Internal split | 0.99 |
| Jiang 2026 [60] | China | Automated interpretation | Aspiration mechanism on VFSS | Support vector machine (ML) | Internal split | — |
| Kim 2023 [61] | Rep. Korea | Automated interpretation | PSD classification (voice) | Multi-branch CNN (DL) | Internal split | 0.95 |
| Ryu 2024 [62] | Rep. Korea | Automated interpretation | Aspiration on VFSS (hyoid) | U-Net tracking (DL) | Apparent only | 0.72 |
| Saab 2023 [63] | Canada | Narrative | Dysphagia screen (voice) | DenseNet/ConvNeXt (DL) | Temporal | 0.91‡ |
| Park 2022 [64] | Rep. Korea | Narrative | Tube-feeding dependence | XGBoost multimodal (ML) | Unclear | 0.85 |
| Hata 2026 [65] | Japan | Narrative | Dysphagia severity (FILS) | LLGMN network (ML) | Cross-validation | — |
| Barron 2024 [66] | USA | Narrative | Aspiration (ultrasound) | VGG16-LSTM (DL) | Internal split | κ only |
| Okamoto 2026 [67] | Japan | Narrative | FILS thresholds / swallowing status | Regularized logistic regression (Reg) | Nested cross-validation | 0.82 |
| Yoon 2024 [68] | Rep. Korea | Narrative | Severe PSD / non-oral feeding | Connectome SVM (ML) | LOOCV | — |

‡The original study reported an invalid 95% CI upper bound exceeding 1.0, rendering the variance uncalculable for pooling. All 14 studies were at high overall risk of bias. VFSS, videofluoroscopic swallowing study; FILS, Food Intake LEVEL Scale; CNN, convolutional neural network; LLGMN, log-linearised Gaussian mixture network; LOOCV, leave-one-out cross-validation; other abbreviations as in Table 1. “—”, no AUC/C-statistic reported.

### 3.3 Primary analysis: early or incident post-stroke dysphagia

The prespecified primary analysis included four studies evaluating models for early or incident PSD. The pooled AUC was 0.94 (95% CI 0.60–0.99), with substantial between-study heterogeneity (I² = 95.6%, τ² = 1.57; Q(3) = 23.6, P < 0.0001) (Figure 2; Table 3).

**Figure 2.**
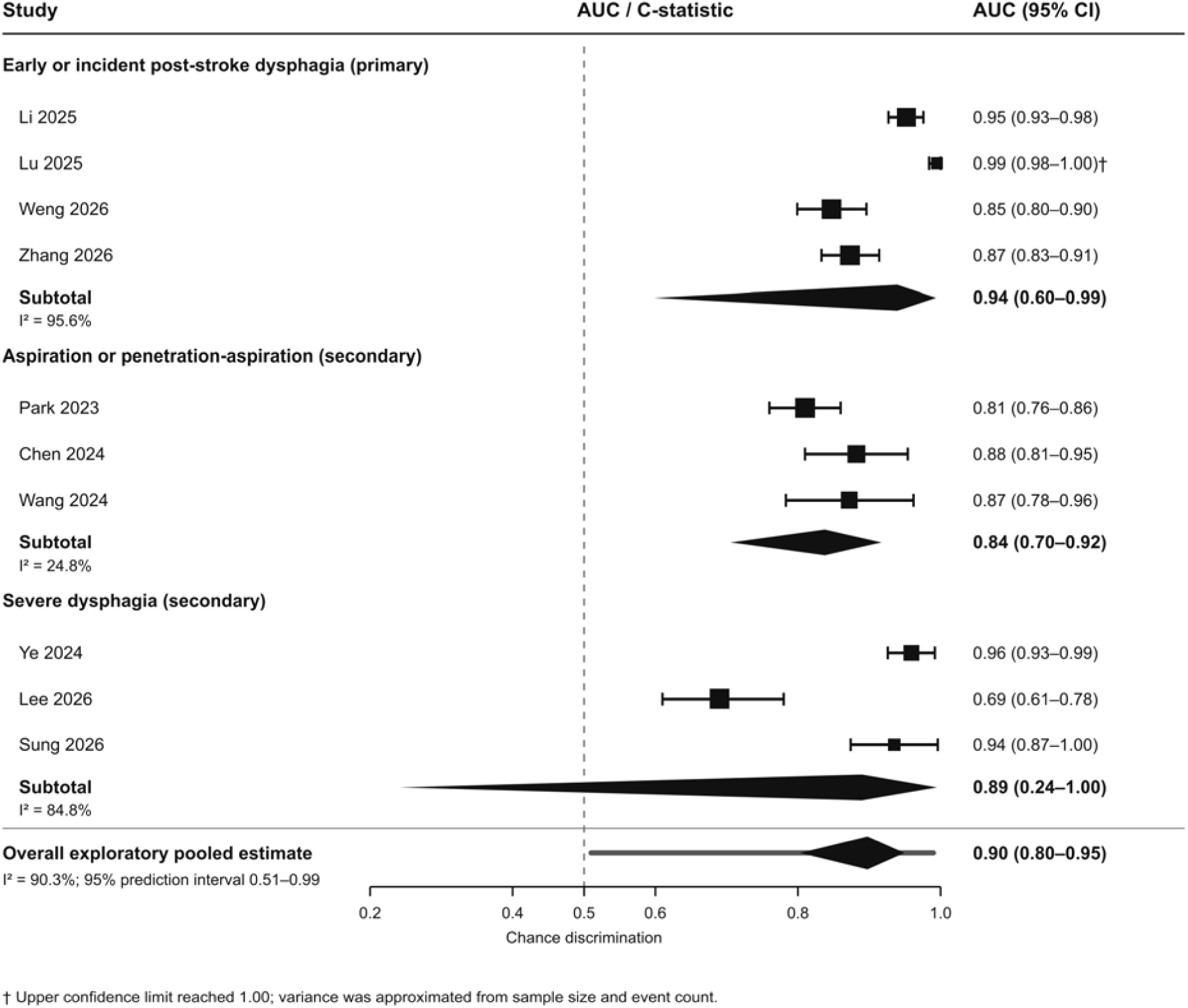
Forest plot of model discrimination (AUC/C-statistic) grouped by outcome stratum. Early or incident post-stroke dysphagia is the prespecified primary analysis; aspiration or penetration-aspiration and severe dysphagia are secondary strata. Squares represent individual study estimates, horizontal lines represent 95% confidence intervals and diamonds represent pooled estimates. The lower diamond summarizes all ten eligible models across outcomes as an exploratory analysis; the thick horizontal line shows its 95% prediction interval (0.51–0.99).

**Table 3.** Pooled model discrimination: pre-registered primary analysis, secondary strata, overall pool and sensitivity analysis.

| Analysis | k | Pooled AUC (95% CI) | I<sup>2</sup> | $\tau^2$ | Q P-value |
| --- | --- | --- | --- | --- | --- |
| Early/incident PSD — PRIMARY | 4 | 0.94 (0.60–0.99) | 95.6% | 1.57 | <0.0001 |
| Aspiration/PA (secondary) | 3 | 0.84 (0.71–0.92) | 24.8% | 0.04 | 0.33 |
| Severe dysphagia (secondary) | 3 | 0.89 (0.24–1.00) | 84.8% | 1.54 | <0.001 |
| Overall — all eligible models (secondary) | 10 | 0.90 (0.80–0.95) | 90.3% | 0.75 | <0.0001 |
| Sensitivity: CI-derived SE, excl. AUC ≥ 0.99 | 9 | 0.88 (0.80–0.93) | 86.0% | 0.44 | <0.0001 |

Random-effects meta-analysis (restricted maximum likelihood) with Hartung-Knapp adjustment, on logit-transformed AUC and back-transformed for presentation. The pre-registered primary analysis was early/incident PSD; the overall pool across outcomes and the other strata are secondary. The overall pool is a field-level exploratory summary and should not be read as evidence that the clinical endpoints are interchangeable. The overall pool had a 95% prediction interval of 0.51-0.99. Descriptive subgroup pooled AUCs were 0.91 for machine-learning models (four studies), 0.92 for regression/nomogram models (five studies), and 0.69 for deep-learning models (one study). By validation design, pooled AUCs were 0.94 for external/temporal validation (four studies) and 0.86 for internal/resampling validation (six studies). These contrasts are descriptive. tau-squared, between-study variance on the logit scale.

The wide confidence interval reflects both the small number of studies and the marked heterogeneity among them. This stratum also included a nomogram with a boundary-level AUC of 0.995. The pooled estimate should therefore be interpreted as highly imprecise despite its high point estimate.

### 3.4 Secondary outcome strata and overall discrimination

In the prespecified secondary analyses, the pooled AUC was 0.84 (95% CI 0.71–0.92; I² = 24.8%) for aspiration or penetration-aspiration (k = 3) and 0.89 (95% CI 0.24–1.00; I² = 84.8%) for severe dysphagia (k = 3) (Figure 2; Table 3).

In the secondary exploratory analysis, pooling all ten eligible clinical risk-prediction models regardless of the specific outcome yielded an AUC of 0.90 (95% CI 0.80–0.95). Heterogeneity was high (I² = 90.3%, τ² = 0.75; Q(9) = 61.7, P < 0.0001), and individual AUC estimates ranged from 0.69 to 0.995 (Figure 2).

The 95% prediction interval for the overall analysis ranged from 0.51 to 0.99, indicating that the discrimination observed in a new setting could plausibly range from little better than chance to near-perfect. Because this analysis combined models addressing different clinical outcomes, it should be interpreted only as a broad summary of reported discrimination and not as evidence that the outcomes are clinically interchangeable.

### 3.5 Subgroup analyses

Pooled discrimination was similar for machine-learning models (AUC 0.91, k = 4) and data-driven regression models or nomograms (AUC 0.92, k = 5). The only deep-learning model, an imaging-based vision transformer developed for lateral medullary infarction, had an AUC of 0.69.

Models evaluated using external or temporal validation had a pooled AUC of 0.94 (k = 4), compared with 0.86 (k = 6) for models evaluated only through internal validation or resampling. This finding should not be interpreted as evidence that external validation improves model performance. Given the small number of studies, sparse subgroup categories and influence of a boundary-level estimate, these subgroup findings are descriptive only.

### 3.6 Sensitivity and robustness analyses

The pooled estimate from the overall analysis was stable in leave-one-out analyses, ranging from 0.876 to 0.909 depending on which study was excluded. The largest decrease occurred after removal of the temporally validated nomogram with a boundary-level AUC of 0.995 (Supplementary Table S2).

Restricting the analysis to studies with variances derived from reported confidence intervals and excluding the boundary-level estimate yielded a pooled AUC of 0.88 (95% CI 0.80–0.93; I² = 86.0%; k = 9) (Table 3).

Small-study effects were not formally evaluated because the overall analysis contained only ten studies with heterogeneous outcomes, model classes and validation designs.

### 3.7 Risk of bias, applicability and reporting

All 24 studies were judged to be at high overall risk of bias using PROBAST or PROBAST+AI. The analysis domain was rated as high risk in every study. Common concerns included small development samples and limited numbers of outcome events, reliance on internal validation, infrequent or incomplete assessment of calibration, and possible data leakage in several acoustic and automated instrumental-interpretation models that used same-event or non-patient-independent data partitions.

External validation was reported in a minority of studies, and calibration was reported in approximately one quarter. Availability of study data, code and complete model specifications was inconsistent.

Applicability concerns were common when the model outcome, such as feeding-tube dependence or automated interpretation of a swallowing assessment, differed from the intended clinical task of predicting early PSD. Readiness was classified as exploratory for most models; a few were considered candidates for further validation, but none met the evidence threshold for clinical implementation. Domain-level assessments for all 24 studies are provided in Supplementary Table S1.

### 3.8 Narrative and secondary synthesis

The 14 studies not included in the meta-analysis were grouped into three categories. Five prognostic or recovery models predicted swallowing recovery, prolonged dysphagia or feeding-tube requirement and reported AUCs ranging from 0.79 to 0.99. Three automated interpretation models classified aspiration or dysphagia directly using VFSS or acoustic data.

The remaining six studies evaluated voice or acoustic signals, ultrasound, structural connectomics or probabilistic networks. These studies were synthesized narratively because they did not report a poolable AUC, used validation methods that were not independent at the patient level, or addressed outcomes outside the prespecified meta-analytic strata.

Although several studies reported high apparent performance, the same risk-of-bias and applicability concerns affected the meta-analyzed studies. None combined independent external validation with adequate assessment of calibration.

## 4. Discussion

### 4.1 Principal findings

The central finding was the variability in reported performance rather than a single summary AUC. The primary estimate for early or incident PSD had a high point estimate but a wide confidence interval, and the prediction interval for the exploratory analysis of all ten models extended from 0.51 to 0.99. Because those models addressed different clinical targets, the overall estimate describes the published literature rather than the performance expected from a model introduced into a new stroke service. Uniformly high risk of bias and sparse calibration or independent validation further reduce confidence that the reported discrimination would transport to routine care.

### 4.2 Interpreting reported model performance

Discrimination was similar between machine-learning models and regression-based models or nomograms. This finding is consistent with broader evidence showing no consistent performance advantage of flexible algorithms over logistic regression when applied to structured, relatively low-dimensional clinical data, particularly when sample sizes and event counts are limited.[28–29] Many of the included models were developed using small samples with few outcome events relative to the number of candidate predictors, increasing the risk of overfitting and optimistic performance estimates.[30] Data leakage was a particular concern in several acoustic and automated instrumental-interpretation studies, especially when swallowing events from the same recordings or patients were distributed across development and evaluation datasets.[31] Inadequate handling of missing data and the use of class-imbalance corrections such as oversampling may introduce additional bias and impair calibration without necessarily improving clinically useful discrimination.[14,32]

High discrimination does not by itself establish clinical validity. A model may rank patients correctly while producing inaccurate absolute risk estimates, and calibration was reported in only a minority of the included studies.[33–34] External validation was also uncommon, although independent evaluation should precede clinical implementation and often produces lower performance estimates than those observed during model development.[35–36] The higher pooled AUC observed among externally or temporally validated models should therefore not be interpreted as evidence of superior transportability. More plausible explanations include selective reporting of successful validation results, small-study optimism, sparse subgroup counts and the influence of a boundary-level estimate. Additional heterogeneity likely arose from differences in dysphagia definitions, assessment timing and reference standards, which also complicate outcome ascertainment in related stroke complications such as stroke-associated pneumonia.[2,37] A concurrent systematic review of aspiration-risk prediction models in stroke reached a similar conclusion, reporting AUCs ranging from 0.76 to 0.96 alongside uniformly high risk of bias.[13]

Reproducibility and assessment of equity were also limited. Availability of code, complete model specifications and supporting data was inconsistent, although these elements are important for independent validation and reconstruction of individual predictions.[16,38] Performance by sex, gender and other clinically relevant subgroups was rarely reported, preventing assessment of differential performance across patient populations. Post hoc explanation methods also have important limitations and should not substitute for transparent model development, complete reporting and independent validation.[39–40]

### 4.3 Clinical implications

Current guidelines recommend swallowing screening before oral intake in patients with acute stroke. Prediction models should therefore be considered potential adjuncts to routine screening rather than replacements for established clinical assessment.[6,41] A sufficiently validated model could help prioritize speech-language pathology referral or instrumental assessment, support decisions regarding oral intake and nutritional route, and identify patients who may benefit from additional airway or pneumonia-prevention measures.[6–7] Clinical value would depend on successful integration into workflow and, ultimately, on impact studies demonstrating that model-guided care changes clinical decisions and improves patient outcomes rather than merely predicting them.[42–43]

### 4.4 Limitations and research priorities

The evidence base also places practical limits on these conclusions. Only ten studies could be pooled, and their clinical targets and designs were not fully comparable. One variance estimate required approximation with the Hanley–McNeil method, and the sparse reporting of calibration, decision curves and net benefit prevented quantitative synthesis beyond discrimination. Most cohorts came from East Asia, leaving performance in other populations and care pathways uncertain. Individual participant data were unavailable, and the screening approach all included studies and quantitative data rather than fully independent duplicate screening of every record—may have left some selection errors undetected. Prospective registration, a broad search without language restrictions and prediction-model-specific appraisal frameworks made the review process transparent, but they cannot remove limitations in the primary studies. Future work should use adequately sized development cohorts, standardized outcomes, internal-external and prospective multicenter validation, complete model reporting and formal impact evaluation.[26,44]

## 5. Conclusion

Reported discrimination was often high, but the evidence does not yet establish transportable clinical performance. Heterogeneous outcomes, uniformly high risk of bias, and limited calibration and independent validation preclude recommending any model for routine care. Future studies should use prespecified outcome definitions, adequately sized development cohorts, independent multicenter validation and prospective evaluation of calibration, clinical utility and impact.

## Ethics

Ethical approval was not required because this systematic review and meta-analysis used previously published data and involved no new human or animal participants.

## Data availability

The data-extraction workbook, analysis-ready dataset, R analysis code and meta-analytic outputs are available from the corresponding author on reasonable request.

