## Supplementary Methods S1 for "Machine learning and data-driven models for predicting post-stroke dysphagia: a systematic review and meta-analysis"

**Supplementary Methods S1. Search strategy**

Five databases were searched from inception through 7 June 2026, with no language restriction: PubMed/MEDLINE, Embase, Web of Science Core Collection, CINAHL (via EBSCO) and the Cochrane Central Register of Controlled Trials (CENTRAL). No full-text, publication-type or peer-reviewed filters were applied except where required by a platform. The exact strings run in each database are reproduced verbatim below.

**A1. PubMed / MEDLINE**

( "Stroke"[Mesh] OR "Brain Ischemia"[Mesh] OR "Intracranial Hemorrhages"[Mesh] OR stroke*[tiab] OR poststroke[tiab] OR "post-stroke"[tiab] OR "cerebrovascular accident*"[tiab] OR "brain infarct*"[tiab] OR "cerebral infarct*"[tiab] OR "ischemic stroke"[tiab] OR "ischaemic stroke"[tiab] OR "intracerebral hemorrhag*"[tiab] OR "intracerebral haemorrhag*"[tiab] OR "subarachnoid hemorrhag*"[tiab] OR "subarachnoid haemorrhag*"[tiab] ) AND ( "Deglutition Disorders"[Mesh] OR dysphagia[tiab] OR "swallowing disorder*"[tiab] OR "deglutition disorder*"[tiab] OR "impaired swallowing"[tiab] OR "swallowing dysfunction"[tiab] OR aspiration[tiab] OR "silent aspiration"[tiab] OR "penetration aspiration"[tiab] OR "swallowing recovery"[tiab] OR "feeding tube"[tiab] OR "tube feeding"[tiab] OR "oral intake"[tiab] ) AND ( "Artificial Intelligence"[Mesh] OR "Machine Learning"[Mesh] OR "Deep Learning"[Mesh] OR "Neural Networks, Computer"[Mesh] OR "artificial intelligence"[tiab] OR "machine learning"[tiab] OR "deep learning"[tiab] OR "neural network*"[tiab] OR "support vector"[tiab] OR "random forest"[tiab] OR "decision tree*"[tiab] OR "gradient boosting"[tiab] OR XGBoost[tiab] OR LightGBM[tiab] OR CatBoost[tiab] OR AdaBoost[tiab] OR "k-nearest neighbor*"[tiab] OR "naive Bayes"[tiab] OR "ensemble model*"[tiab] OR radiomic*[tiab] OR algorithm*[tiab] OR "natural language processing"[tiab] OR "automated model*"[tiab] ) AND ( predict*[tiab] OR prognos*[tiab] OR "risk model*"[tiab] OR "prediction model*"[tiab] OR "predictive model*"[tiab] OR "classification model*"[tiab] OR "risk stratification"[tiab] OR "model development"[tiab] OR validation[tiab] ) NOT ( animals[mh] NOT humans[mh] )

**A2. Embase (Elsevier)**

( 'stroke'/exp OR 'brain ischemia'/exp OR 'intracranial hemorrhage'/exp OR stroke*:ti,ab OR poststroke:ti,ab OR 'post-stroke':ti,ab OR 'cerebrovascular accident*':ti,ab OR 'brain infarct*':ti,ab OR 'cerebral infarct*':ti,ab OR 'ischemic stroke':ti,ab OR 'ischaemic stroke':ti,ab OR 'intracerebral hemorrhag*':ti,ab OR 'intracerebral haemorrhag*':ti,ab OR 'subarachnoid hemorrhag*':ti,ab OR 'subarachnoid haemorrhag*':ti,ab ) AND ( 'dysphagia'/exp OR 'deglutition disorder'/exp OR dysphagia:ti,ab OR 'swallowing disorder*':ti,ab OR 'deglutition disorder*':ti,ab OR 'impaired swallowing':ti,ab OR 'swallowing dysfunction':ti,ab OR aspiration:ti,ab OR 'silent aspiration':ti,ab OR 'penetration aspiration':ti,ab OR 'swallowing recovery':ti,ab OR 'feeding tube':ti,ab OR 'tube feeding':ti,ab OR 'oral intake':ti,ab ) AND ( 'artificial intelligence'/exp OR 'machine learning'/exp OR 'deep learning'/exp OR 'neural network'/exp OR 'artificial intelligence':ti,ab OR 'machine learning':ti,ab OR 'deep learning':ti,ab OR 'neural network*':ti,ab OR 'support vector':ti,ab OR 'random forest':ti,ab OR 'decision tree*':ti,ab OR 'gradient boosting':ti,ab OR xgboost:ti,ab OR lightgbm:ti,ab OR catboost:ti,ab OR adaboost:ti,ab OR 'k-nearest neighbor*':ti,ab OR 'naive bayes':ti,ab OR 'ensemble model*':ti,ab OR radiomic*:ti,ab OR algorithm*:ti,ab OR 'natural language processing':ti,ab OR 'automated model*':ti,ab ) AND ( predict*:ti,ab OR prognos*:ti,ab OR 'risk model*':ti,ab OR 'prediction model*':ti,ab OR 'predictive model*':ti,ab OR 'classification model*':ti,ab OR 'risk stratification':ti,ab OR 'model development':ti,ab OR validation:ti,ab ) AND ( [humans]/lim ) NOT ( 'animal'/exp NOT 'human'/exp )

**A3. Web of Science Core Collection**

TS=( stroke* OR poststroke OR "post-stroke" OR "cerebrovascular accident*" OR "brain infarct*" OR "cerebral infarct*" OR "ischemic stroke" OR "ischaemic stroke" OR "intracerebral hemorrhag*" OR "intracerebral haemorrhag*" OR "subarachnoid hemorrhag*" OR "subarachnoid haemorrhag*" ) AND TS=( dysphagia OR "swallowing disorder*" OR "deglutition disorder*" OR "impaired swallowing" OR "swallowing dysfunction" OR aspiration OR "silent aspiration" OR "penetration aspiration" OR "swallowing recovery" OR "feeding tube" OR "tube feeding" OR "oral intake" ) AND TS=( "artificial intelligence" OR "machine learning" OR "deep learning" OR "neural network*" OR "support vector" OR "random forest" OR "decision tree*" OR "gradient boosting" OR XGBoost OR LightGBM OR CatBoost OR AdaBoost OR "k-nearest neighbor*" OR "naive Bayes" OR "ensemble model*" OR radiomic* OR algorithm* OR "natural language processing" OR "automated model*" ) AND TS=( predict* OR prognos* OR "risk model*" OR "prediction model*" OR "predictive model*" OR "classification model*" OR "risk stratification" OR "model development" OR validation )

**A4. CINAHL via EBSCOhost**

Run each concept as a separate row using TX All Text, then combine S1 AND S2 AND S3 AND S4.

S1 stroke* OR poststroke OR "post-stroke" OR "cerebrovascular accident*" OR "brain infarct*" OR "cerebral infarct*" OR "ischemic stroke" OR "ischaemic stroke" OR "intracerebral hemorrhag*" OR "intracerebral haemorrhag*" OR "subarachnoid hemorrhag*" OR "subarachnoid haemorrhag*"

S2 dysphagia OR "swallowing disorder*" OR "deglutition disorder*" OR "impaired swallowing" OR "swallowing dysfunction" OR aspiration OR "silent aspiration" OR "penetration aspiration" OR "swallowing recovery" OR "feeding tube" OR "tube feeding" OR "oral intake"

S3 "artificial intelligence" OR "machine learning" OR "deep learning" OR "neural network*" OR "support vector" OR "random forest" OR "decision tree*" OR "gradient boosting" OR XGBoost OR LightGBM OR CatBoost OR AdaBoost OR "k-nearest neighbor*" OR "naive Bayes" OR "ensemble model*" OR radiomic* OR algorithm* OR "natural language processing" OR "automated model*"

S4 predict* OR prognos* OR "risk model*" OR "prediction model*" OR "predictive model*" OR "classification model*" OR "risk stratification" OR "model development" OR validation

S5 S1 AND S2 AND S3 AND S4

**A5. Cochrane CENTRAL (Search Manager)**

#1 stroke* OR poststroke OR post NEXT stroke OR cerebrovascular NEXT accident* OR ischemic NEXT stroke OR ischaemic NEXT stroke OR brain NEXT infarct* OR cerebral NEXT infarct* OR intracerebral NEXT hemorrhag* OR intracerebral NEXT haemorrhag* OR subarachnoid NEXT hemorrhag* OR subarachnoid NEXT haemorrhag*
#2 dysphagia OR swallowing OR deglutition OR aspiration OR swallowing NEXT disorder* OR deglutition NEXT disorder* OR impaired NEXT swallowing OR swallowing NEXT dysfunction OR silent NEXT aspiration OR penetration NEXT aspiration OR swallowing NEXT recovery OR feeding NEXT tube OR tube NEXT feeding OR oral NEXT intake
#3 artificial NEXT intelligence OR machine NEXT learning OR deep NEXT learning OR neural NEXT network* OR support NEXT vector OR random NEXT forest OR decision NEXT tree* OR gradient NEXT boosting OR XGBoost OR LightGBM OR CatBoost OR AdaBoost OR k NEXT nearest NEXT neighbor* OR naive NEXT Bayes OR ensemble NEXT model* OR radiomic* OR algorithm* OR natural NEXT language NEXT processing OR automated NEXT model*
#4 predict* OR prognos* OR validation OR risk NEXT model* OR prediction NEXT model* OR predictive NEXT model* OR classification NEXT model* OR risk NEXT stratification OR model NEXT development
#5 #1 AND #2 AND #3 AND #4
