## Supplementary material for "Machine learning and data-driven models for predicting post-stroke dysphagia: a systematic review and meta-analysis": Table S1

**Table S1. Domain-level risk of bias for all 24 included studies.**

| **Study** | **Participants** | **Predictors** | **Outcome** | **Analysis** | **Overall** |
| --- | --- | --- | --- | --- | --- |
| Park 2023 | Unclear | Low | Low | High | High |
| Ye 2024 | High | Unclear | High | High | High |
| Wang 2024 | Low | High | High | High | High |
| Chen 2024 | High | Low | High | High | High |
| Li 2025 | Unclear | Low | Unclear | High | High |
| Lu 2025 | High | High | Unclear | High | High |
| Sung 2026 | High | Low | High | High | High |
| Zhang 2026 | High | Low | High | High | High |
| Weng 2026 | High | Unclear | Low | High | High |
| Lee 2026 | High | Low | Unclear | High | High |
| Lee 2020 | High | Low | Unclear | High | High |
| Tian 2026 | High | Low | Low | High | High |
| Ito 2021 | Unclear | Low | Unclear | High | High |
| Zhang 2022 | High | Unclear | Unclear | High | High |
| Seo 2025 | High | Low | Unclear | High | High |
| Jiang 2026 | High | High | Low | High | High |
| Kim 2023 | High | High | Unclear | High | High |
| Ryu 2024 | High | High | Unclear | High | High |
| Saab 2023 | High | Unclear | High | High | High |
| Park 2022 | High | Unclear | Unclear | High | High |
| Hata 2026 | High | Unclear | High | High | High |
| Barron 2024 | High | Low | Low | High | High |
| Okamoto 2024 | High | High | Unclear | High | High |
| Yoon 2024 | High | Low | Unclear | High | High |

Assessed with PROBAST and, for AI/ML models, PROBAST+AI. The analysis domain was rated high risk in every study, driving the high overall rating throughout. Low, low risk of bias; High, high risk of bias; Unclear, insufficient information.
