## Supplementary material for "Machine learning and data-driven models for predicting post-stroke dysphagia: a systematic review and meta-analysis": Table S2

**Table S2. Leave-one-out analysis for the exploratory overall AUC meta-analysis.**

| **Study omitted** | **Pooled AUC** | **95% CI** |
| --- | --- | --- |
| Park 2023 | 0.907 | 0.806–0.959 |
| Ye 2024 | 0.888 | 0.777–0.947 |
| Wang 2024 | 0.902 | 0.792–0.957 |
| Chen 2024 | 0.902 | 0.790–0.957 |
| Li 2025 | 0.886 | 0.772–0.947 |
| Lu 2025 | 0.876 | 0.800–0.926 |
| Sung 2026 | 0.895 | 0.784–0.953 |
| Zhang 2026 | 0.903 | 0.792–0.958 |
| Weng 2026 | 0.905 | 0.798–0.959 |
| Lee 2026 | 0.909 | 0.829–0.954 |

Random-effects model (restricted maximum likelihood with Hartung–Knapp adjustment) on logit-transformed AUC. The pooled estimate remained between 0.876 and 0.909 across all omissions; the largest reduction followed removal of Lu 2025, the boundary-flagged nomogram (AUC 0.995).
