## Supplementary material for "Machine learning and data-driven models for predicting post-stroke dysphagia: a systematic review and meta-analysis": Table S3

**Table S3. Study-level input for the primary meta-analyses and exploratory overall analysis.**

| **Study** | **Outcome stratum** | **Model class** | **Validation** | **N (events)** | **AUC (95% CI)** | **SE, logit AUC** | **Variance source** |
| --- | --- | --- | --- | --- | --- | --- | --- |
| Park 2023 | Aspiration/PA | Machine learning | Random split-sample | 3408 (448) | 0.810 (0.760–0.860) | 0.169 | Reported 95% CI |
| Ye 2024 | Severe dysphagia | Machine learning | Independent external cohort | 724 (308) | 0.959 (0.926–0.992) | 0.585 | Reported 95% CI |
| Wang 2024 | Aspiration/PA | Regression/nomogram | Geographic external validation | 528 (89) | 0.872 (0.783–0.962) | 0.497 | Reported 95% CI |
| Chen 2024 | Aspiration/PA | Regression/nomogram | Temporal validation | 412 (184) | 0.882 (0.810–0.954) | 0.404 | Reported 95% CI |
| Li 2025 | Early/incident PSD | Machine learning | Random split-sample | 1041 (305) | 0.952 (0.927–0.976) | 0.297 | Reported 95% CI |
| Lu 2025 | Early/incident PSD | Regression/nomogram | Temporal validation | 250 (100) | 0.995 (0.984–1.000)† | 1.008 | Hanley–McNeil approximation |
| Sung 2026 | Severe dysphagia | Regression/nomogram | Random split-sample | 212 (66) | 0.935 (0.874–0.996) | 0.913 | Reported 95% CI |
| Zhang 2026 | Early/incident PSD | Machine learning | Random split-sample | 908 (464) | 0.873 (0.833–0.914) | 0.193 | Reported 95% CI |
| Weng 2026 | Early/incident PSD | Regression/nomogram | Bootstrap | 394 (79) | 0.847 (0.799–0.896) | 0.197 | Reported 95% CI |
| Lee 2026 | Severe dysphagia | Deep learning | Random split-sample | 163 (44) | 0.690 (0.610–0.780) | 0.209 | Reported 95% CI |

AUC estimates were transformed to the logit scale for analysis. Standard errors were derived from reported 95% confidence intervals except for Lu 2025. † Upper confidence limit reached 1.00; the variance was approximated using the Hanley–McNeil method from the sample size and number of events. PA, penetration-aspiration.
