## Supplementary material for "Machine learning and data-driven models for predicting post-stroke dysphagia: a systematic review and meta-analysis": Table S4

**Table S4. Reports excluded after full-text assessment and primary reason for exclusion.**

| **Report** | **Primary reason for exclusion** |
| --- | --- |
| Pan Z. et al. (2024). Machine-learning Assisted Swallowing Assessment: A Deep Learning-based Qualit.... Neurology. https://doi.org/10.1212/wnl.0000000000208174. | Duplicate/overlapping dataset |
| Perry L (2001). Screening swallowing function of patients with acute stroke. Part one: identi.... Journal of Clinical Nursing (Wiley-Blackwell). https://doi.org/10.1046/j.1365-2702.2001.00501.x. | No AI/ML or data-driven model |
| Oto T, Kandori Y, Ohta T, Domen K, Koyama T. (2009). Predicting the chance of weaning dysphagic stroke patients from enteral nutri.... Eur J Phys Rehabil Med. | No AI/ML or data-driven model |
| Wang Z, Shi Y, Zhang L, Wu L, Fang Q, Huiling L. (2022). Nomogram for predicting swallowing recovery in patients after dysphagic stroke. JPEN J Parenter Enteral Nutr. https://doi.org/10.1002/jpen.2115. | No AI/ML or data-driven model |
| Hu X. et al. (2025). Predictors and lesion patterns of dysphagia and swallowing outcomes after acu.... Therapeutic Advances in Neurological Disorders. https://doi.org/10.1177/17562864241311130. | No AI/ML or data-driven model |
| Uchimura M. et al. (2026). Decoupling of swallowing and motor recovery after aneurysmal subarachnoid hem.... Neurochirurgie. https://doi.org/10.1016/j.neuchi.2026.101794. | No AI/ML or data-driven model |
| Phan T.G. et al. (2019). Stroke Severity Versus Dysphagia Screen as Driver for Post-stroke Pneumonia. Frontiers in Neurology. https://doi.org/10.3389/fneur.2019.00016. | No dysphagia/swallowing outcome |
| John Paul II Hospital, Krakow (2021). Acute Stroke of CArotid Artery Bifurcation Origin Treated With Use oF The Mic.... clinicaltrials.gov. | No dysphagia/swallowing outcome |
| Deng YM, Sun JJ, Gu HQ, Yang KX, Wang YJ, Li ZX, Zhao XQ. (2024). Predictors of dysphagia screening and pneumonia among patients with intracere.... BMJ Open. https://doi.org/10.1136/bmjopen-2023-073977. | No dysphagia/swallowing outcome |
| Liang Y, Dai X, Wei B, Jia H, Zhang J, Qiu Z, Zhang Q. (2025). Development and Validation of a GULP-Based Predictive Model for Dehydration i.... Br J Hosp Med (Lond). https://doi.org/10.12968/hmed.2024.0366. | No dysphagia/swallowing outcome |
| Han A, Zhang S, Yao Y, He J, Wang Y, Yang H. (2025). Development and validation of a risk predication model for nutritional risk b.... PLoS One. https://doi.org/10.1371/journal.pone.0330982. | No dysphagia/swallowing outcome |
| Styczen H. et al. (2025). Impact of imaging biomarkers from body composition analysis on outcome of end.... Journal of NeuroInterventional Surgery. https://doi.org/10.1136/jnis-2024-022275. | No dysphagia/swallowing outcome |
| Nishioka S. et al. (2017). Malnutrition risk predicts recovery of full oral intake among older adult str.... Clinical Nutrition. https://doi.org/10.1016/j.clnu.2016.06.028. | No extractable performance metric |
| Jijakli A. et al. (2024). Quality Improvement Initiative Using Predictive Swallowing Score to Guide Nut.... Neurology: Clinical Practice. https://doi.org/10.1212/cpj.0000000000200352. | No extractable performance metric |
| Runions S et al. (2004). Practice on an acute stroke unit after implementation of a decision-making al.... Journal of Neuroscience Nursing. https://doi.org/10.1097/01376517-200408000-00006. | No multivariable prediction model |
| George MG et al. (2009). Paul Coverdell National Acute Stroke Registry Surveillance -- four states, 20.... MMWR Surveillance Summaries. | No multivariable prediction model |
| Martino R et al. (2009). The Toronto Bedside Swallowing Screening Test (TOR-BSST): development and val.... Stroke (00392499). https://doi.org/10.1161/strokeaha.107.510370. | No multivariable prediction model |
| Okubo P.C.M.I. et al. (2012). Using the national institute of health stroke scale to predict dysphagia in a.... Cerebrovascular Diseases. https://doi.org/10.1159/000336240. | No multivariable prediction model |
| Umay, EK et al. (2013). Evaluation of Dysphagia in Early Stroke Patients by Bedside, Endoscopic, and .... DYSPHAGIA. https://doi.org/10.1007/s00455-013-9447-z. | No multivariable prediction model |
| Marcella Rachadel Avelino et al. (2017). Oral dietary intake level in thrombolysed and non-thrombolysed patients after ischemic stroke. NeuroRehabilitation. https://doi.org/10.3233/nre-161389. | No multivariable prediction model |
| Carnaby, Giselle et al. (2019). Associations Between Spontaneous Swallowing Frequency at Admission, Dysphagia.... Archives of Physical Medicine & Rehabilitation. https://doi.org/10.1016/j.apmr.2019.01.009. | No multivariable prediction model |
| Cuellar, Megan E. et al. (2019). Predictive value of laryngeal adductor reflex testing in patients with dyspha.... International Journal of Speech-Language Pathology. https://doi.org/10.1080/17549507.2018.1512652. | No multivariable prediction model |
| Ng K.B. et al. (2021). Classification of Stroke Patients With Dysphagia Into Subgroups Based on Patt.... Archives of Physical Medicine and Rehabilitation. https://doi.org/10.1016/j.apmr.2020.11.014. | No multivariable prediction model |
| Liang J, Yin Z, Li Z, Gu H, Yang K, Xiong Y, Wang Y, Wang C. (2022). Predictors of dysphagia screening and pneumonia among patients with acute isc.... Stroke Vasc Neurol. https://doi.org/10.1136/svn-2020-000746. | No multivariable prediction model |
| Pereira, Vitor Costa et al. (2023). Post-stroke dysphagia: Clinical characteristics and evolution in a single-pri.... NeuroRehabilitation. https://doi.org/10.3233/nre-220242. | No multivariable prediction model |
| Horn, Janet et al. (2024). The Relationship Between Poststroke Dysphagia and Poststroke Depression and I.... American Journal of Speech-Language Pathology. https://doi.org/10.1044/2024_ajslp-23-00264. | No multivariable prediction model |
| Horn, Janet et al. (2026). An Exploratory Classification Framework for Poststroke Dysphagia Severity Usi.... American Journal of Speech-Language Pathology. https://doi.org/10.1044/2025_ajslp-24-00313. | No multivariable prediction model |
| Moss, M et al. (2020). Development of an Accurate Bedside Swallowing Evaluation Decision Tree Algori.... CHEST. https://doi.org/10.1016/j.chest.2020.07.051. | Not adult human stroke population |
| Lee, JT et al. (2020). Machine learning analysis to automatically measure response time of pharyngea.... SCIENTIFIC REPORTS. https://doi.org/10.1038/s41598-020-71713-4. | Not adult human stroke population |
| Jauk, S et al. (2023). Evaluation of a Machine Learning-Based Dysphagia Prediction Tool in Clinical .... DYSPHAGIA. https://doi.org/10.1007/s00455-022-10548-9. | Not adult human stroke population |
| Vaitheeshwari, R et al. (2023). The Swallowing Intelligent Assessment System Based on Tongue Strength and Surface EMG. IEEE SENSORS JOURNAL. https://doi.org/10.1109/jsen.2023.3277825. | Not adult human stroke population |
| Mazzeo, S et al. (2024). Dysphagia in primary progressive aphasia: Clinical predictors and neuroanatom.... EUROPEAN JOURNAL OF NEUROLOGY. https://doi.org/10.1111/ene.16370. | Not adult human stroke population |
| Song, HD et al. (2025). Development and validation of a screening model for dysphagia in the elderly .... FRONTIERS IN MEDICINE. https://doi.org/10.3389/fmed.2025.1719174. | Not adult human stroke population |
| Liu, S et al. (2025). Identify predictive factors for the emergence of self-reported oropharyngeal .... FRONTIERS IN NEUROLOGY. https://doi.org/10.3389/fneur.2025.1439579. | Not adult human stroke population |
| Jeong CW, Lim DW, Noh SH, Moon HK, Park C, Ko N, Kim MS. (2025). Multi-Center Validation of Artificial Intelligence-Based Video Analysis Platf.... Diagnostics (Basel). https://doi.org/10.3390/diagnostics16010045. | Not adult human stroke population |
| Balci, S et al. (2025). Non-routine thrombectomy in pediatric arterial ischemic stroke. DIAGNOSTIC AND INTERVENTIONAL RADIOLOGY. https://doi.org/10.4274/dir.2024.242675. | Not adult human stroke population |
| Dai, YN et al. (2026). Interpretable machine learning for accessible dysphagia screening and staging.... ISCIENCE. https://doi.org/10.1016/j.isci.2025.114451. | Not adult human stroke population |
| Gao Y.-F. et al. (2023). Identification of influencing factors and development of a nomogram for nutri.... World Chinese Journal of Digestology. https://doi.org/10.11569/wcjd.v31.i21.904. | Outcome not post-stroke dysphagia-related |
| 李雅楠 et al. (2024). 基于机器学习算法构建缺血性脑卒中伴吞咽障碍 患者卒中相关性肺炎模型并验证. Chinese Journal of Convalescent Medicine / Zhongguo Liaoyang Yixue. https://doi.org/10.13517/j.cnki.ccm.2024.10.001. | Outcome not post-stroke dysphagia-related |
| Bao L. et al. (2025). Immunoinflammatory biomarkers as predictors of hemorrhagic transformation in .... Frontiers in Neurology. https://doi.org/10.3389/fneur.2025.1606563. | Outcome not post-stroke dysphagia-related |
| Zeng F. et al. (2026). Development and validation of the nomogram of aspiration pneumonia in stroke .... Medicine (United States). https://doi.org/10.1097/md.0000000000047875. | Outcome not post-stroke dysphagia-related |
| García Gálvez J.A. et al. (2026). Febrile complications in acute stroke: can we anticipate them from admission?. Revista Cientifica de la Sociedad Espanola de Enfermeria Neurologica. https://doi.org/10.1016/j.sedene.2026.500208. | Outcome not post-stroke dysphagia-related |
| 赵佳月 et al. (2026). 老年脑卒中病人口腔衰弱变化轨迹 及影响因素.. Chinese Nursing Research. https://doi.org/10.12102/j.issn.1009-6493.2026.04.006. | Outcome not post-stroke dysphagia-related |
| Furtner J. (2023). Craniofacial musculature assessment: A novel technique for predicting stroke .... European Journal of Radiology. https://doi.org/10.1016/j.ejrad.2023.111024. | Protocol/review/editorial/commentary |
| Wang, C et al. (2026). Postoperative Rebleeding: The Sword of Damocles in Minimally Invasive Surgery.... RESEARCH. https://doi.org/10.34133/research.1083. | Protocol/review/editorial/commentary |
| Huhmann M et al. (2004). Comparison of dysphagia screening by a registered dietitian in acute stroke p.... Topics in Clinical Nutrition. https://doi.org/10.1097/00008486-200407000-00008. | Stroke-specific data not separable |
| Kritas, S et al. (2016). Objective prediction of pharyngeal swallow dysfunction in dysphagia through a.... NEUROGASTROENTEROLOGY AND MOTILITY. https://doi.org/10.1111/nmo.12730. | Stroke-specific data not separable |
| Catriona M. Steele et al. (2019). Development of a Non-invasive Device for Swallow Screening in Patients at Risk of Oropharyngeal Dysphagia: Results from a Prospective Exploratory Study. Dysphagia. https://doi.org/10.1007/s00455-018-09974-5. | Stroke-specific data not separable |
| Roldan-Vasco, S et al. (2021). Machine learning based analysis of speech dimensions in functional oropharyng.... COMPUTER METHODS AND PROGRAMS IN BIOMEDICINE. https://doi.org/10.1016/j.cmpb.2021.106248. | Stroke-specific data not separable |
| Chang-Won Jeong et al. (2024). The Development of an Artificial Intelligence Video Analysis-Based Web Application to Diagnose Oropharyngeal Dysphagia: A Pilot Study. Brain Sciences. https://doi.org/10.3390/brainsci14060546. | Stroke-specific data not separable |
| Zhang B, Wong KP, Liu M, Hui V, Guo C, Liu Z, Liu Y, Xiao Q, Qin J. (2025). Effect of artificial intelligence-based video-game system on dysphagia in pat.... Clin Nutr. https://doi.org/10.1016/j.clnu.2024.12.022. | Treatment effect only |

The three reports that could not be retrieved are shown separately in Figure 1 and are not included in this table. The primary reason follows the review's prespecified eligibility hierarchy; a report may have met more than one exclusion criterion.
